# Association of PNPLA3 rs738409 G/C gene polymorphism with nonalcoholic fatty liver disease in children: a meta-analysis

**DOI:** 10.1101/2020.05.19.20106385

**Authors:** Shan Tang, Jing Zhang, Ting-Ting Mei, Hai-Qing Guo, Xin-Huan Wei, Wen-Yan Zhang, Ya-Li Liu, Shan Liang, Zuo-Peng Fan, Li-Xia Ma, Wei Lin, Yi-Rong Liu, Li-Xia Qiu, Hai-Bin Yu

**Affiliations:** Department of Hepatitis C and drug-induced liver injury, Beijing YouAn Hospital, Capital Medical University, Beijing 100069 China

**Author notes:** Correspondence to Dr HaiBin Yu.

## Abstract

**Objective:** To systematically review the association of PNPLA3 rs738409 G/C gene polymorphism with non-alcoholic fatty liver disease(NAFLD) in children.

**Design:** Systematic review and meta-analysis.

**Data sources and study selection:** We searched MEDLINE, PubMed, EMBASE, and CENTRAL databases from inception to May 2019. Case-control studies assessing the relationship between PNPLA3 rs738409 G/C gene polymorphism with non-alcoholic fatty liver disease in children were selected according to inclusion and exclusion criteria.

**Data extraction and analyses:** First author’s surname, publication year, country, total numbers of patients in the case and control groups, sex ratio, and body mass index (BMI), as well as the numbers of cases and controls with the C/C, C/G and G/G genotypes were extracted from the included studies. Random effects model was used to quantify the association between the PNPLA3 rs738409 G/C gene polymorphism and the susceptibility of children’s NAFLD. Fixed effects model was used to quantify the relationship between the PNPLA3 rs738409 G/C gene polymorphism and the severity of NAFLD in children.

**Results:** A total of nine case-control studies were included in this meta-analysis containing data of 1173 children with NAFLD and 1792 healthy controls. Five studies compared NAFLD children and non-NAFLD healthy populations. Statistical analysis showed that PNPLA3 gene polymorphism was significantly associated with children’s NAFLD in the allele contrast, dominant, recessive and over dominant models (G vs C,OR=3.343, 95% CI=1.524-7.334; GG+GC vs CC,OR=3.157, 95% CI=1.446-6.892;GG vs GC+CC,OR=5.692, 95% CI=1.941-16.689; GG+CC vs GC,OR=2.756, 95% CI=1.729-4.392). Four case-control studies compared Children with nonalcoholic fatty liver (NAFL) and children with nonalcoholic steatohepatitis (NASH). The results showed that the PNPLA3 gene polymorphism was also significantly associated with the severity of NAFLD in children in recessive gene model(GG vs GC+CC,OR = 14.43, 95% CI = 5.985-34.997); The Egger’s test revealed no significant publication bias.

**Conclusions:** Meta-analysis showed that PNPLA3 gene polymorphism was significantly associated with susceptibility and severity of NAFLD in children.

**trial registration number:** CRD42019134056.

**strengths and limitations of this study:** 1.Published and previously unavailable data were synthesised to estimate the association of PNPLA3 rs738409 G/C gene polymorphism with NAFLD in children.2. This meta-analysis not only explored the relationship between PNPLA3 gene polymorphism and susceptibility of NAFLD in children, but also researched the relationship between PNPLA3 gene polymorphism and susceptibility of NASH in children. It can be further investigated whether PNPLA3 gene can be used to early diagnosis children NAFLD and evaluate the severity of NAFLD in children.3. High between-study heterogeneity and the likelihood of publication bias may impact the generalisability of our results.4. This meta-analysis only involved single factor studies, the interaction of PNPLA3 gene polymorphism and obesity,breastfeeding time were not taken into consideration.

## INTRODUCTION

Nonalcoholic fatty liver disease (NAFLD) is one of the most common causes of chronic liver disease worldwide. The prevalence of NAFLD in children has reached about 10%, and in obese children has reached 40%-70%^1^.NAFLD includes nonalcoholic fatty liver (NAFL), nonalcoholic steatohepatitis (NASH),fatty liver fibrosis and cirrhosis. NAFL usually has no obvious clinical symptoms, but hepatocyte injury can lead to the occurrence of NASH, liver fibrosis and cirrhosis in 3%-5% of patients^2^. Therefore, assessing the genetic factors of children’s NAFLD to early diagnose the disease, early judge the severity of the disease is the key to improve prognosis. Current studies have shown that PNPLA3 (Patatin-like phospholipase domain containing 3) rs738409 G/C gene polymorphism is associated with adult nonalcoholic fatty liver disease^3,4^. But there is no consensus on the relationship between PNPLA3 rs738409 G/C gene polymorphism and children NAFLD due to differences in population samples, detection methods and diagnostic criteria. To this end, a meta-analysis of published research is conducted to comprehensively assess the relationship between PNPLA3 gene polymorphism and NAFLD in children.

## METHODS

The search strategy, eligibility criteria and outcomes were described a priori (PROSPERO CRD42019134056).

### Data sources

We searched MEDLINE, PubMed, EMBASE and CENTRAL databases for eligible articles without language restriction, from inception to May 2019.

### study selection

We included case control studies that fulfilled the following inclusion criteria:1) the study cohorts include PNPLA3 rs738409 G/C gene polymorphism in children with NAFLD and non-NAFLD individuals;2) the articles clearly stated the diagnostic criteria for NAFLD;3) case-control studies were enrolled, the case group was NAFLD children, the control group was non-NAFLD individuals or the case group was NASH children, and the control group was NAFL children;4) if duplicate research reports was retrieved, the most comprehensive one was selected to avoid repeated statistics;5) The full text can be retrieved. Articles were excluded based on the following criteria: 1) the source of the enrolled cases in the literature is unclear;2) the enrolled case groups are not children patients;3) the data collection and analysis methods in the literature are unscientific; 4) the OR values and 95% CI were not available by calculation; 5) non-case-control studies; 6) animal studies.

### Data extraction

Two reviewers independently extracted data from the original publications: author, year of publication, country of origin, design of the trial, total numbers of patients in the case and control groups, sex ratio, body mass index (BMI), the numbers of cases and controls with the C/C, C/G, and G/G genotypes, whether genotype distribution was consistent with the Hardy-Weinberg equilibrium (HWE). The selected literature was extracted by two researchers and encountered controversial issues, experts were invited to review.

### Data analysis

The association of the G/C polymorphism in the PNPLA3 gene with NAFLD in children susceptibility was evaluated in the allelic, dominant, recessive, and super-dominant models. The association of the G/C polymorphism in the PNPLA3 gene with NASH in children susceptibility was evaluated in the recessive model. Meta-analysis statistical treatments were performed using Stata 12.0 software, and the results were evaluated using the OR value and the corresponding 95% confidence interval (CI). Heterogeneity across studies was determined by the Q-and *I*^2^ tests. The fixed-effects model was used in the case of nonsignificant heterogeneity (*P*>0.05, *I*^2^<50%); otherwise, the random-effects model was utilized. The suitability test was used to check whether the gene distribution of the control group consistent with the Hardy-Weinberg equilibrium, and the P_H-W_ >0.05 was regarded as consistent with the HWE. The sensitivity analysis was performed by excluding individual documents sequentially to evaluate its effect on the overall results under all genetic models. The publication bias was evaluated using the Egger linear regression method.

## RESULTS

### search results

According to the set search formula, 267 articles that meet the requirements were initially retrieved, and 123 duplicate articles were removed. 82 unrelated documents were excluded after reading the title and abstract.Then 53 articles were excluded after reading the full text leaving 9 articles that were included in the final analysis. The literature inclusion process is shown in Figure 1.

**Figure 1.**
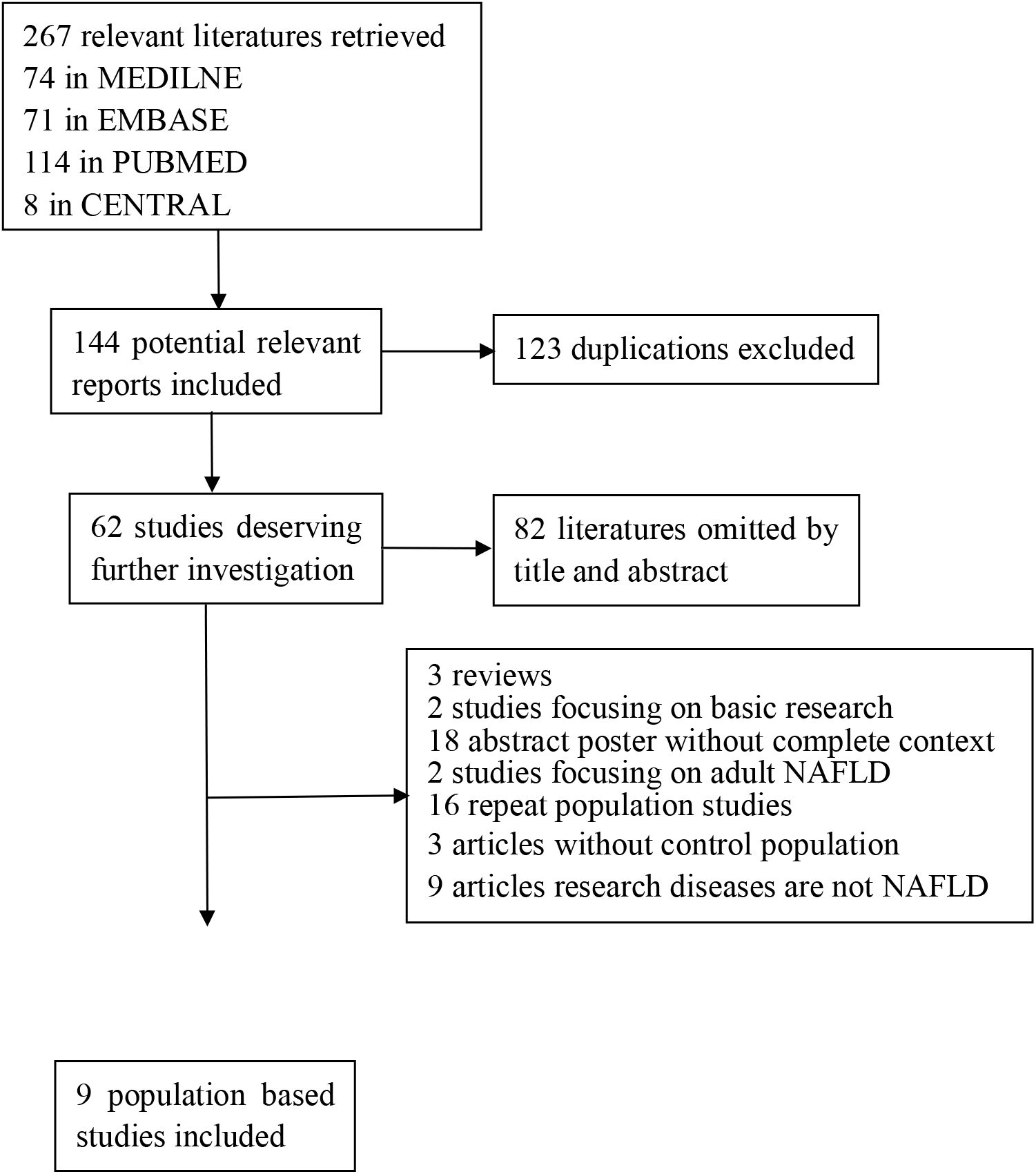
Flow diagram for study selection.

### Characteristics of the included studies

Table 1 depicts the basic features of all five included studies. A total of 1173 children with NAFLD and 1792 individuals without NAFLD were included. Genotyping data for all studies are summarized in Table 1.

**Table 1.**
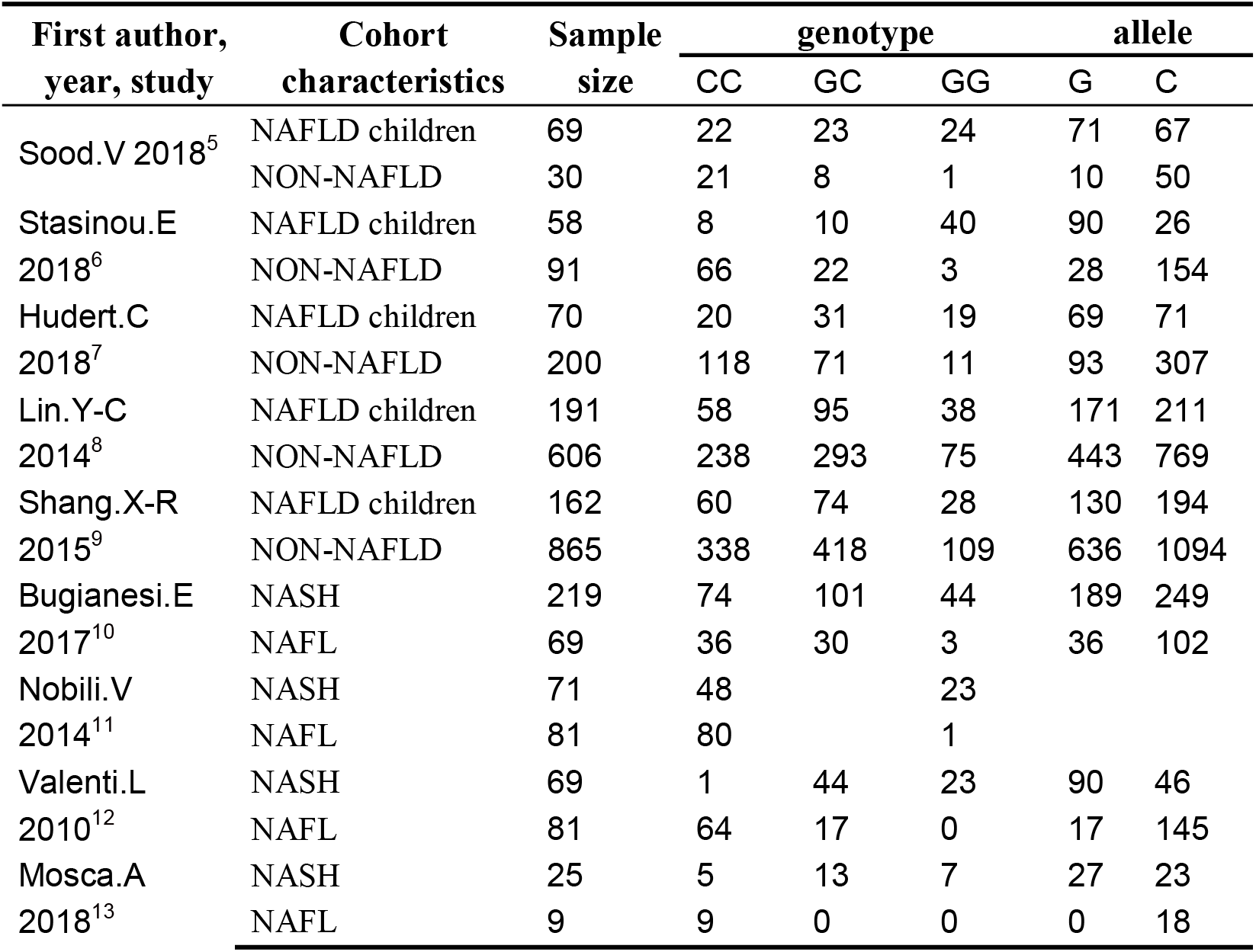
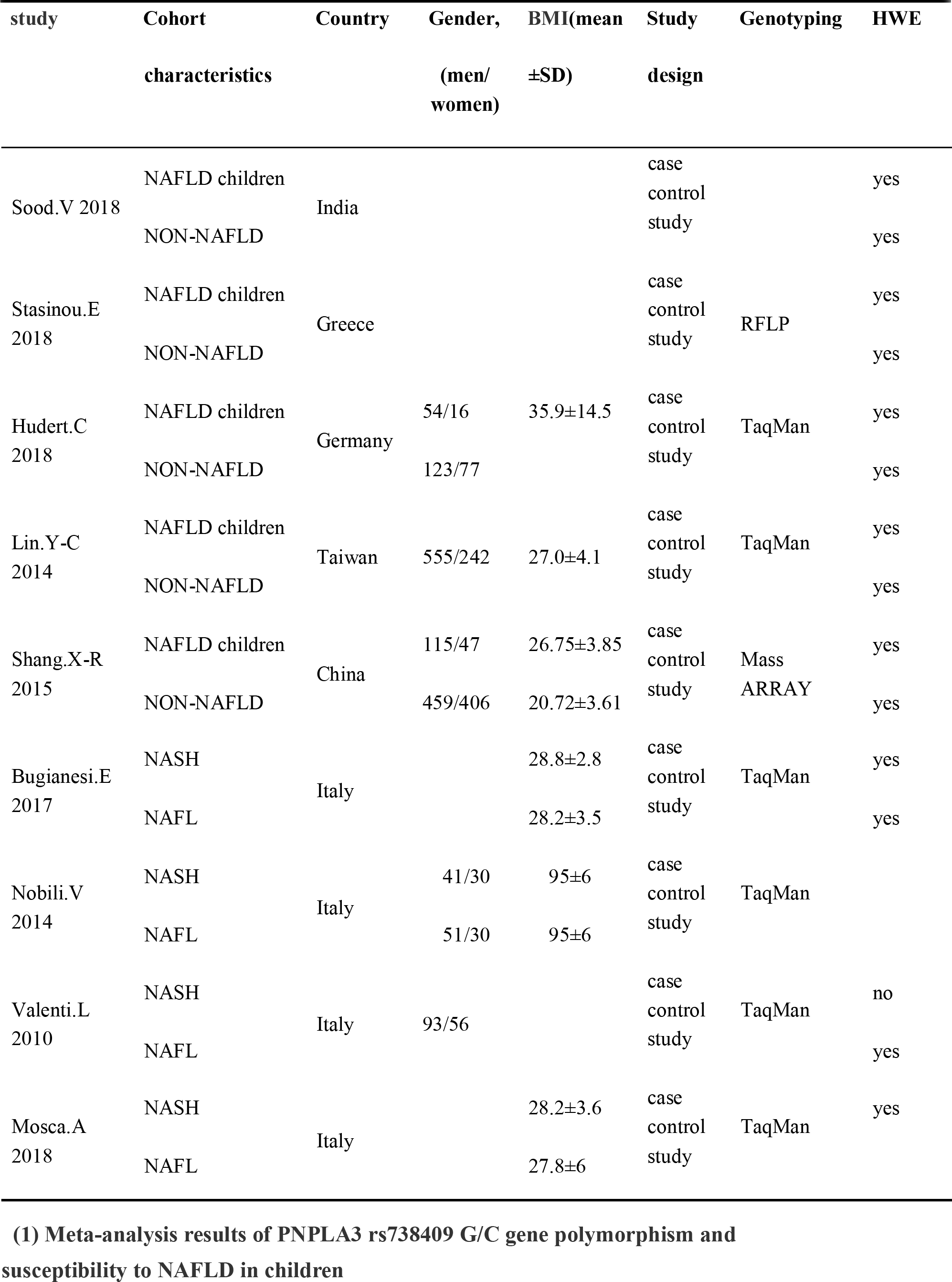
Characteristics of the studies included in the meta-analysis.

### Meta-analysis results

A total of 9 articles included in the current meta-analysis. 5 literatures described the association between PNPLA3 rs738409 G/C gene polymorphism and susceptibility to childhood NAFLD disease. The allelic (G vs C), dominant (GC+GG vs CC), recessive (GG vs CG+CC), and super-dominant (CC+GG vs GC) models were performed in this meta-analysis. There were 550 children with NAFLD and 1792 non-NAFLD controls. The random effects model was employed for pooled ORs since significant heterogeneity was detected. Meta-analysis showed that the PNPLA3 gene polymorphism was significantly associated with susceptibility to NAFLD in children.

4 articles described the association between PNPLA3 rs738409 G/C gene polymorphism and the severity of NAFLD in children. A recessive gene model (GG vs CG+CC) analysis was performed in this meta analysis. There were 383 children with NASH as case group and 240 children with NAFL control as control group contained in this meta analysis. In the heterotypic analysis, the heterogeneity of each study was nonsignificant, the fixed effect model was used to combine the OR values. Meta-analysis showed that the PNPLA3 gene polymorphism was significantly associated with the severity of NAFLD in children.

**(1) Meta-analysis results of PNPLA3 rs738409 G/C gene polymorphism and susceptibility to NAFLD in children**

**PNPLA3 rs738409 G/C in the dominant gene model (GG+GC vs CC):**

GG+GC genotypes was used as exposure factor and CC genotype as non-exposure factor for analysis. There were 382 cases with GG+GC genotypes and 168 cases with CC genotypes in the case group. Non-NAFLD individuals as control group had 1011 GG+GC genotypes and 781 CC genotypes. The results showed that the risk of NAFLD was higher in GG and GC genotypes than in CC genotype. (GG+GC vs CC OR=3.157, 95% CI=1.446-6.892, P=0.004; figure 2)

**Figure 2.**
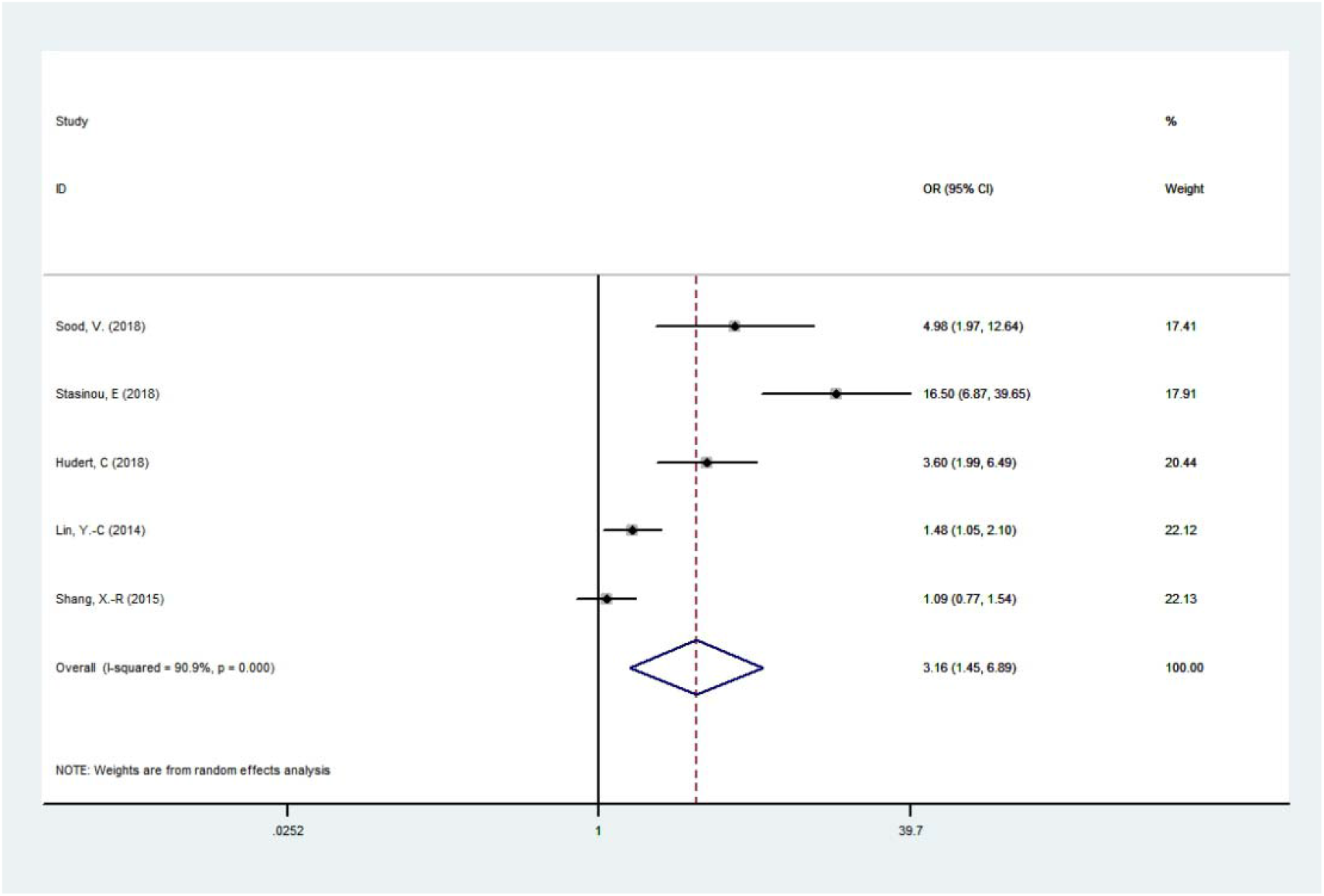
Forest plot of studies evaluating the OR with 95%CI of PNPLA3 rs738409 G/C in the dominant model(GG+GC vs CC) in NAFLD children. CI, Confidence interval; OR, odds ratio

**PNPLA3 rs738409 G/C in the allele model (G vs C):**

The G allele as the exposure factor and the C allele as the non-exposure factor were analyzed. The combined analysis showed 531 G alleles and 569 C alleles in the case group. The control group had 1210 G alleles and 2374 C alleles. Mata analysis result showed that PNPLA3 rs738409 G/C polymorphism was significantly correlated with NAFLD in children. (G vs C OR=3.343, 95% CI=1.524-7.334, P=0.003; figure 3).

**Figure 3.**
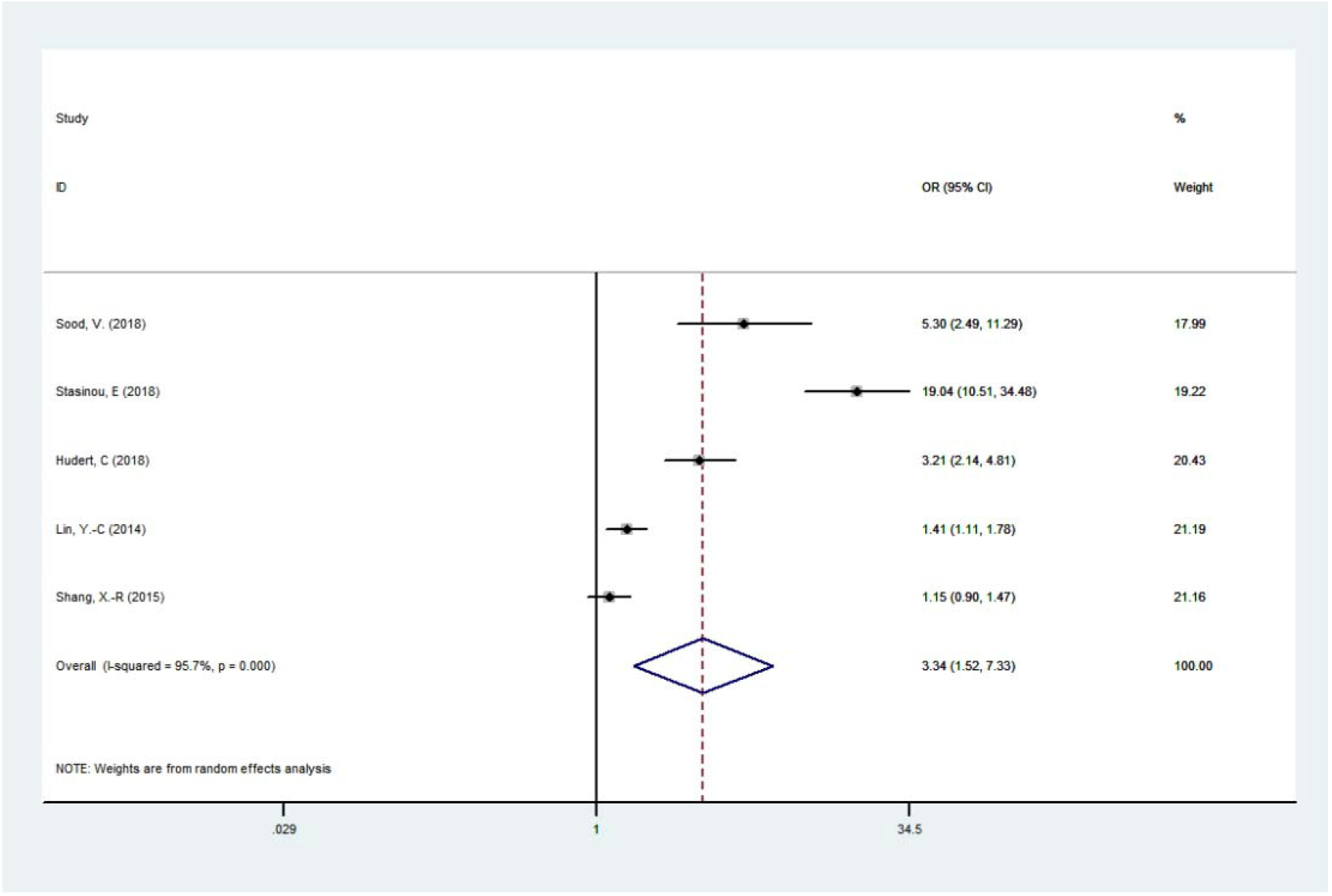
Forest plot of studies evaluating the OR with 95%CI of PNPLA3 rs738409 G/C in the allele model (G vs C) in NAFLD children. CI, Confidence interval; OR, odds ratio

**PNPLA3 rs738409 G/C in the recessive gene model (GG vs CG+CC):**

GG genotype used as exposure factor and GC+CC genotypes as non-exposure factor were analyzed. A total of 149 children had the GG genotype and 1530 displayed the GC+CC genotypes among cases. Meanwhile, 199 and 1593 cases had the GG and GC+CC genotypes among controls. The results showed that the risk of NAFLD in children with GG genotype was higher than that of GC+CC genotypes. (GG vs GC+CC OR=5.692, 95% CI=1.941-16.689, P=0.002; figure 4).

**Figure 4.**
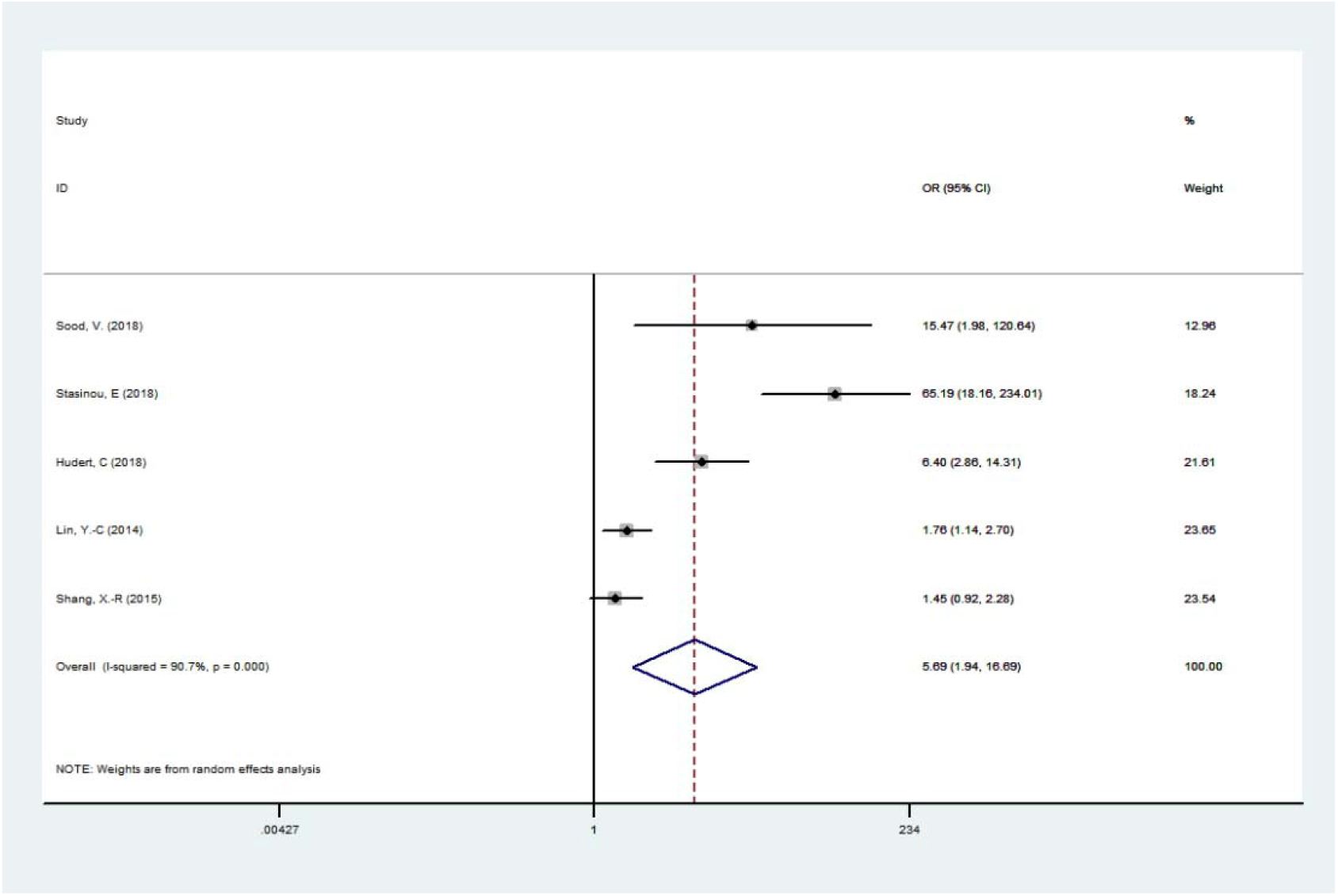
Forest plot of studies evaluating the OR with 95%CI of PNPLA3 rs738409 G/C in the recessive gene model (GG vs CG+CC) in NAFLD children. CI, Confidence interval; OR, odds ratio

**PNPLA3 rs738409 G/C in the superdominant model (GG+CC vs GC):**

GG+CC genotypes was used as the exposure factor and GC genotype as the non-exposure factor. In the case group, there were 1014 GG+CC genotypes and 233 GC genotypes. The control group had 908 GG+CC genotypes and 812 GC genotypes. The results showed that the risk of NAFLD in GG+CC genotypes was higher than that in GC genotype. (GG+CC vs GC OR=2.756, 95% CI=1.729-4.392, P=0.000; figure 5).

**Figure 5.**
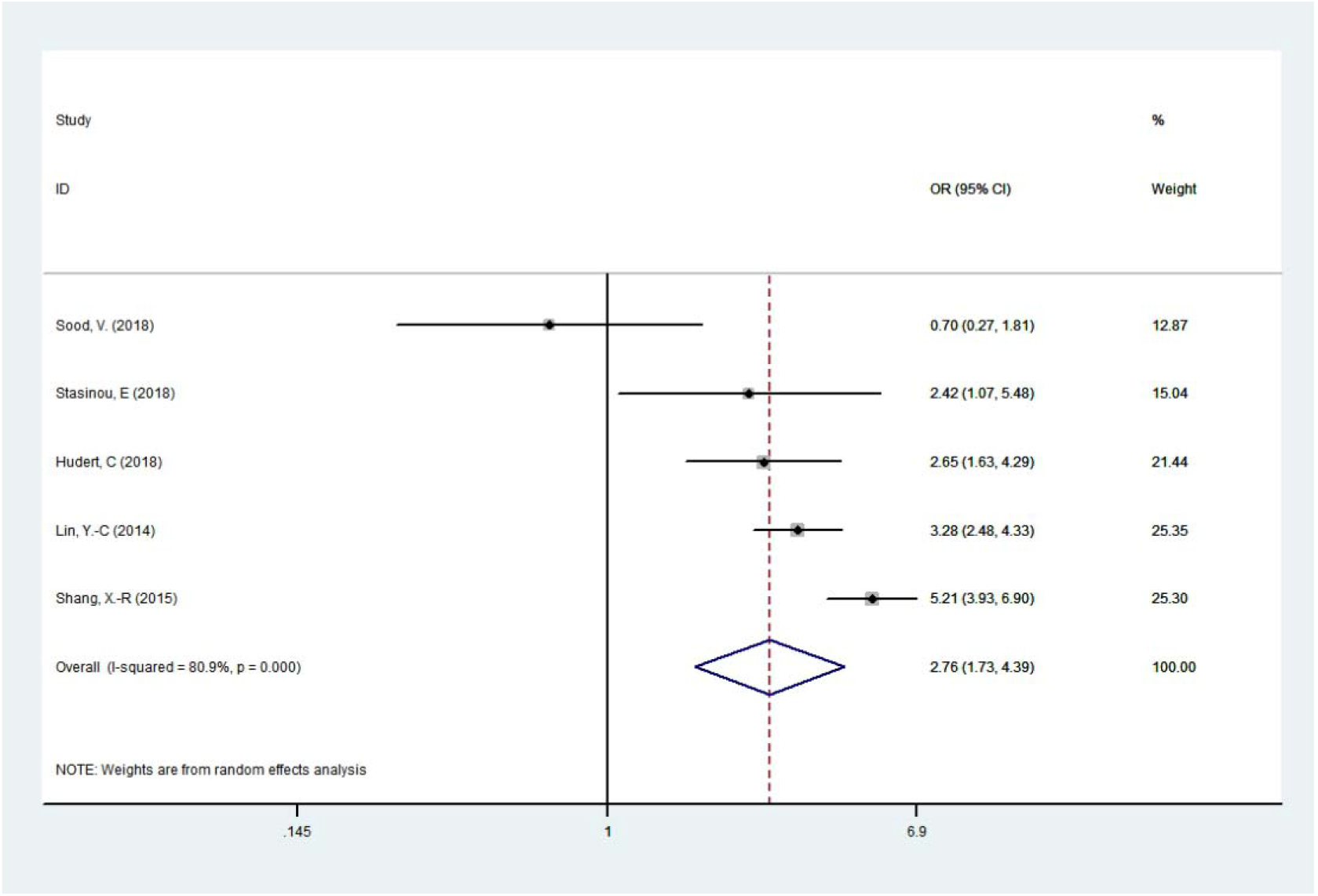
Forest plot of studies evaluating the OR with 95%CI of PNPLA3 rs738409 G/C in the superdominant model (GG+CC vs GC) in NAFLD children. CI, Confidence interval; OR, odds ratio

**(2) Meta-analysis result of PNPLA3 rs738409 G/C gene polymorphism and the severity of NAFLD in children**

**PNPLA3 rs738409 G/C recessive gene model (GG vs CG+CC):**

The GG genotype as the exposure factor and the GC+CC genotype as the non-exposure factor were analyzed. In NASH group, there were 236 GG genotypes and 4 GC+CC genotypes. In NAFL group, there were 286 GG genotypes and 97 GC+CC genotypes. The results showed that the risk of NASH in NAFLD patients with GG genotype was higher than that in GC+CC genotypes. (GG vs GC+CC OR=14.473, 95% CI=5.985-34.997, P=0.000; figure 6).

**Figure 6.**
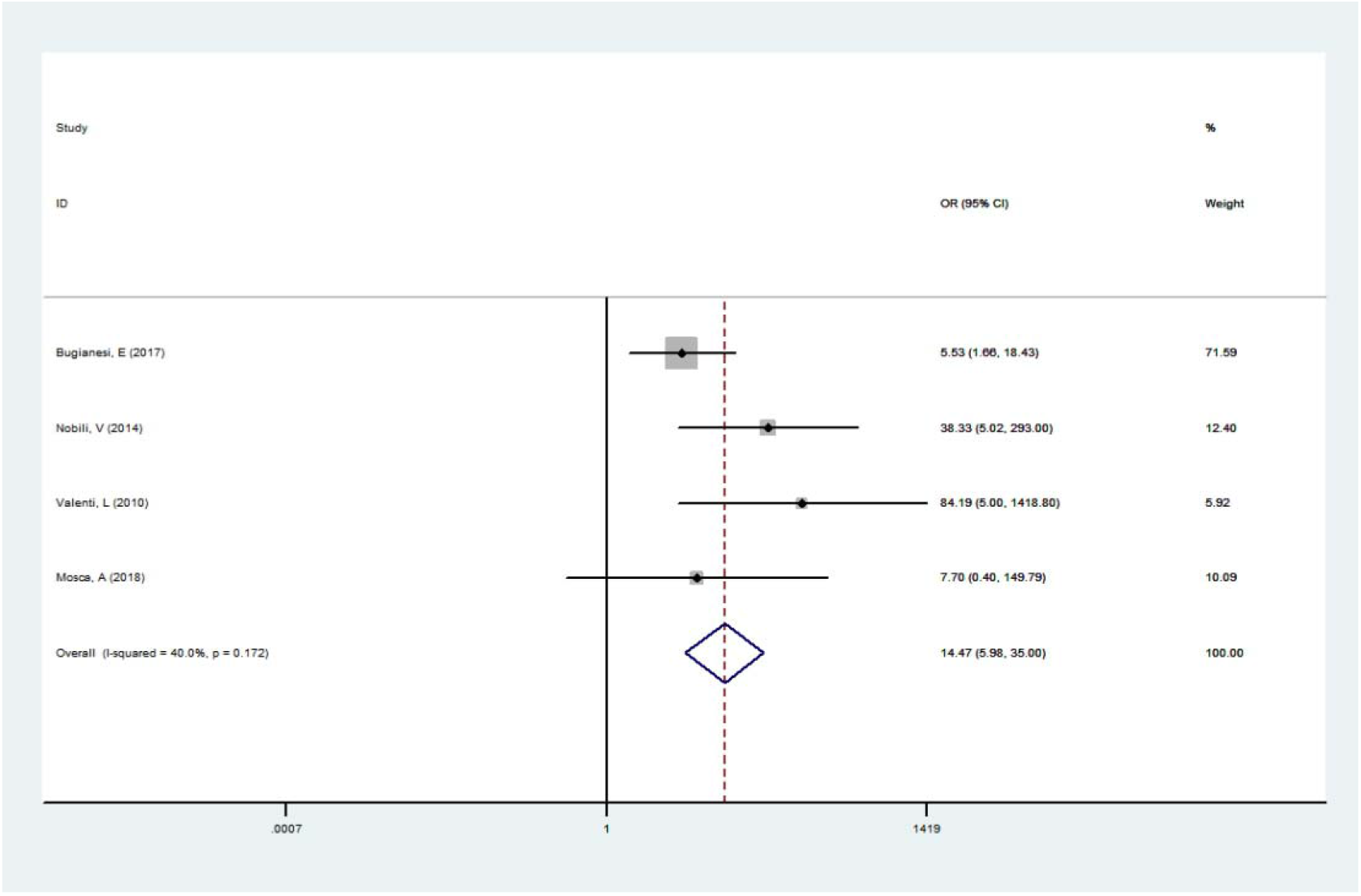
Forest plot of studies evaluating the OR with 95%CI of PNPLA3 rs738409 G/C in the recessive gene model (GG vs CG+CC) in NASH children. CI, Confidence interval; OR, odds ratio

**Table 2.** Meta-analysis of the association of PNPLA3 rs738409 G/C gene polymorphism and childhood NAFLD susceptibility.

| Genetic model | Relevance test |  |  | Heterogeneity test |  |  | Publication |  |
| --- | --- | --- | --- | --- | --- | --- | --- | --- |
|  | OR(95% CI) | Z | P<sub>value</sub> | I<sup>2</sup> | Q | P<sub>het</sub> | P<sub>egger</sub> | t |
| dominant gene model | 3.157 ( 1.446-6.892 ) | 4.51 | 0.000 | 48.5 | 7.38 | 0.117 | 0.060 | 2.95 |
| Allelic model | 3.343 ( 1.524-7.334 ) | 3.01 | 0.003 | 95.7 | 93.77 | 0.000 | 0.081 | 2.59 |
| recessive model | 5.692 ( 1.941-16.689 ) | 3.17 | 0.002 | 90.7 | 42.91 | 0.000 | 0.076 | 2.67 |
| super-dominant model | 2.756 ( 1.729-4.392 ) | 4.26 | 0.000 | 80.9 | 20.93 | 0.000 | 0.038 | -3.56 |

**Table 3.** Meta-analysis of the association of PNPLA3 rs738409 G/C gene polymorphism and childhood NASH susceptibility.

| Genetic model | Relevance test |  |  | Heterogeneity test |  |  | Publication |  |
| --- | --- | --- | --- | --- | --- | --- | --- | --- |
|  | OR(95% CI) | Z | P<sub>value</sub> | I<sup>2</sup> | Q | P<sub>hel</sub> | P<sub>egger</sub> | t |
| recessive model | 14.473 ( 5.985-34.997 ) | 5.93 | 0.000 | 40.0 | 5.00 | 0.172 | 0.602 | 0.61 |

### Sensitivity analysis

In the meta analysis of the relationship betweenPNPLA3 rs738409 G/C gene polymorphism and children NAFLD disease susceptibility, Sensitivity analysis was performed by omitting one study sequentially to examine its effect on the overall results under all genetic models. In the four genetic models of PNPLA3 rs738409 G/C, OR values obtained after eliminating any one of the studies were close to pre-exclusion ORs, indicating the robustness of the current analysis (Supplementary Figures 1, 2, 3, and 4).

In the meta-analysis of the relationship between PNPLA3 rs738409 G/C gene polymorphism and childhood NASH susceptibility, Removing single study one by one method was used to analyze the sensitivity. In the recessive gene model of PNPLA3 rs738409 G/C, the OR value obtained after the elimination of any one of the studies was relatively close to that before the elimination, indicating that the results of this study were robust. (Supplementary Figure 5).

### Publication bias analysis

In the meta-analysis of PNPLA3 rs738409 G/C gene polymorphism and childhood NAFLD susceptibility, the Egger regression analysis of 4 gene models suggested that no publication bias existed, including the allelic (G vs C, P=0.081), dominant (GC+GGvs CC, P=0.060), recessive (GG vs GC+CC, P=0.076) and super-dominant (CC+GG vs GC, P=0.038) models (Supplementary Figures 5, 6, 7, and 8).

In the meta-analysis of PNPLA3 rs738409 G/C gene polymorphism and NAFLD severity in children, the Egger regression analysis of the recessive gene model suggested no publication bias existed(GG vs GC+CC, P=0.172).(Supplementary Figure 9)

## DISCUSSION

The phospholipase domain protein 3(PNPLA3) was originally cloned from the cDNA library of 3T3 preadipocytes into mature adipocytes, hence named adiponectin^14^. It is 10 times more expressed in the liver than in adipose tissue^15^. Non-synonymous genetic variation (rs738409) in the human patatin-like phospholipase domain-containing 3 gene (PNPLA3) that substitutes methionine for isoleucine (I148M) at amino acid position 148 in exon 3 was found to be associated with remodeling of liver triglycerides^16^. Hepatocytes expressing PNPLA3-I148M have more long-chain polyunsaturated fatty acids (PUFA)^17^. Mitsche et al reported that hepatocytes expressing PNPLA3-I148M could transfer PUFA from triglyceride to phospholipid^18^. Phospholipid remodeling is associated with NAFLD susceptibility.

Although the research on the relationship between PNPLA3 gene polymorphism and NAFLD in children has attracted the attention of many researchers, the results vary from study to study. The stability and reliability of the research results of a single study are affected by the small sample size. However, meta-analyses use suitable mathematical models to perform quantitative analysis of multiple identical or similar research results, increasing the test efficiency of research results.

In this study, by developing a retrieval strategy, literature quality evaluation was conducted according to the requirements of the Oxford Critical Appraisal Skill Program (Oxford CASP, 2004)^19^, and literature without clear diagnostic criteria and repeated reports were removed. Finally, 9 literatures meeting the requirements were included for data extraction. 5 literatures studied the relationship between PNPLA3 738409 locus gene polymorphism and NAFLD susceptibility in children. The random effect model was used to conduct combined analysis of the data, and then to evaluate whether the PNPLA3 738409 locus gene polymorphism was associated with NAFLD susceptibility in children. Allele model, dominant gene model, recessive gene model and superdominant gene model of PNPLA3 738409 locus were analyzed in this study, and the results showed that PNPLA3 738409 locus gene polymorphism was significantly associated with NAFLD susceptibility in children. 4 literatures studied the relationship between PNPLA3 738409 locus gene polymorphism and NAFLD severity in children. The fixed effect model was used to conduct combined analysis of the data. Due to data limitations, the recessive gene model of PNPLA3 738409 locus was analyzed, and the results showed that PNPLA3 738409 locus gene polymorphism was significantly associated with NASH in children.

In sensitivity analysis, exclusion of individual studies had no effect on the combined effect size, the meta-analysis findings were relatively stable. Publication bias analysis found no significant publication bias in the meta-analysis, suggesting reliable results.

At the same time, there are still some limitations in this article. First, due to the lack of a unified document quality evaluation standard, the included articles are subjectively selected and evaluated, which may affect the stability of the meta-analysis results. Second, since meta-analysis itself is a retrospective study, there is a degree of bias. Due to these limitations, we still need to expand the sample size to further and systematically evaluate case-control studies. In addition, this meta-analysis only involved single-factor studies, and did not include the interaction of PNPLA3 gene polymorphism with obesity^20^ and breastfeeding^21^ for hours, and the interaction of the above factors may affect the susceptibility of NAFLD in children. In summary, the polymorphism of PNPLA3 738409 locus gene polymorphism is not only related to the susceptibility of NAFLD in children, but also related to its severity. Because NAFLD in children has no specific clinical manifestations, and ultrasound is not sensitive to the diagnosis of NASH, further studies can be conducted to evaluate whether PNPLA3 738409 G/C gene polymorphism can be screened for early diagnosis of childhood NAFLD and early evaluation of NAFLD severity.

## Data Availability

All data referred to in the manuscript are available.

## Author contributions

Tang S and Zhang J contributed equally to this work; Tang S designed and wrote the manuscript that led to the submission; Mei TT, Guo HQ, and Wei XH searched and filtered the literature; Zhang WY, Liu YL and Liang S selected and interpreted the data; Fan ZP, Ma LX, Liu YR, Lin W and Qiu LX revised the manuscript; Yu HB conceived the study, Yu HB was corresponding authors, every author read and approved the final manuscript.

## Patient consent for publication

Not required.

## Data sharing statement

No additional data are available.

## References

1 Clemente MG, Mandato C, Poeta M et al. Pediatric non-alcoholic fatty liver disease: Recent solutions, unresolved issues, and future research directions. World J Gastroenterol 2016; 22: 8078–93.

2 Romeo S, Kozlitina J, Xing C et al. Genetic variation in PNPLA3 confers susceptibility to nonalcoholic fatty liver disease. NAT GENET 2008; 40: 1461–5.

3 Mazo DF, Malta FM, Stefano JT et al. Validation of PNPLA3 polymorphisms as risk factor for NAFLD and liver fibrosis in an admixed population. ANN HEPATOL 2019.

4 Hotta K, Yoneda M, Hyogo H et al. Association of the rs738409 polymorphism in PNPLA3 with liver damage and the development of nonalcoholic fatty liver disease. BMC MED GENET 2010; 11: 172.

5 Sood V, Khanna R, Rawat D et al. Predictive risk factors and transient elastography in pediatric non alcoholic fatty liver disease in Indian population. J PEDIATR GASTR NUTR 2018; 66: 84.

6 Stasinou E, Argyraki M, Lambropoulos A et al. Association of the variants rs738409 and rs2896019 in the palatin-like phospholipase 3 gene (PNPLA3) in Greek children and adolescents with fatty liver disease. J PEDIATR GASTR NUTR 2018; 66: 86.

7 Hudert CA, Selinski S, Rudolph B et al. Genetic determinants of steatosis and fibrosis progression in pediatric non-alcoholic fatty liver disease. Liver international: official journal of the International Association for the Study of the Liver 2018.

8 Lin Y, Chang P, Chang M et al. Genetic variants in GCKR and PNPLA3 confer susceptibility to nonalcoholic fatty liver disease in obese individuals. The American journal of clinical nutrition 2014; 99: 869–74.

9 Shang X, Song J, Liu F et al. GWAS-Identified Common Variants With Nonalcoholic Fatty Liver Disease in Chinese Children. J PEDIATR GASTR NUTR 2015; 60: 669–74.

10 Bugianesi E, Bizzarri C, Rosso C et al. Low Birthweight Increases the Likelihood of Severe Steatosis in Pediatric Non-Alcoholic Fatty Liver Disease. AM J GASTROENTEROL 2017; 112: 1277–86.

11 Nobili V, Donati B, Panera N et al. A 4-polymorphism risk score predicts steatohepatitis in children with nonalcoholic fatty liver disease. J Pediatr Gastroenterol Nutr 2014; 58: 632–6.

12 Valenti L, Alisi A, Galmozzi E et al. I148M patatin-like phospholipase domain-containing 3 gene variant and severity of pediatric nonalcoholic fatty liver disease. HEPATOLOGY 2010; 52: 1274–80.

13 Mosca A, Fintini D, Scorletti E et al. Relationship between non-alcoholic steatohepatitis, PNPLA3 I148M genotype and bone mineral density in adolescents. LIVER INT 2018; 38: 2301–8.

14 Basu RS. PNPLA3-I148M: a problem of plenty in non-alcoholic fatty liver disease. ADIPOCYTE 2019; 8: 201–8.

15 Wilson PA, Gardner SD, Lambie NM et al. Characterization of the human patatin-like phospholipase family. J LIPID RES 2006; 47: 1940–9.

16 Baclig MO, Lozano-Kuhne JP, Mapua CA et al. Genetic variation I148M in patatin-like phospholipase 3 gene and risk of non-alcoholic fatty liver disease among Filipinos. INT J CLIN EXP MED 2014; 7: 2129–36.

17 Ruhanen H, Perttila J, Holtta-Vuori M et al. PNPLA3 mediates hepatocyte triacylglycerol remodeling. J LIPID RES 2014; 55: 739–46.

18 Mitsche MA, Hobbs HH, Cohen JC. Patatin-like phospholipase domain-containing protein 3 promotes transfer of essential fatty acids from triglycerides to phospholipids in hepatic lipid droplets. J BIOL CHEM 2018; 293: 9232.

19 Serou N, Sahota L, Husband AK et al. Systematic review of psychological, emotional and behavioural impacts of surgical incidents on operating theatre staff. BJS Open 2017; 1: 106–13.

20 Kleiner DE, Brunt EM. Nonalcoholic fatty liver disease: pathologic patterns and biopsy evaluation in clinical research. SEMIN LIVER DIS 2012; 32: 3–13.

21 Ayonrinde OT, Oddy WH, Adams LA et al. Infant nutrition and maternal obesity influence the risk of non-alcoholic fatty liver disease in adolescents. J HEPATOL 2017; 67: 568–76.

